# Evaluation of dual-active ingredient long-lasting insecticidal nets against wild resistant malaria vectors in Tanzania: a series of experimental hut trials

**DOI:** 10.64898/2026.07.27.26359071

**Authors:** Jackline L. Martin, Jo Lines, Louisa A. Messenger, Edmond Bernard, Sian E. Clarke, Selina Antony, Thomas S. Churcher, Dominic P. Dee, Yvan G. Fotso-Toguem, Ciara L. Hamilton, Jacklin F. Mosha, Raphael N’Guessan, Mark Rowland, Rosine Z. Wolie, Charles S. Wondji, Alphaxard Manjurano

## Abstract

The effectiveness of malaria vector control is increasingly threatened by widespread insecticide resistance, particularly to pyrethroids. In response, the World Health Organization recommends dual-active ingredients (dual-A.I.) insecticide-treated nets (ITNs) in resistant settings. This study evaluated the efficacy of several dual-A.I. ITNs in two experimental hut trial sites an area with high insecticide resistant malaria vectors in Tanzania with differing vector species composition and bionomics.

Five ITNs were evaluated: a pyrethroid-only net (MAGNet®), three pyrethroid-piperonyl butoxide (PBO) nets (OlysetTM Plus, Veeralin®, and PermaNet® 3.0), one dual-A.I. net (Interceptor® G2; chlorfenapyr + alpha-cypermethrin), and an untreated control net. Mosquitoes were collected from huts, morphologically identified, and monitored for mortality up to 72 h. WHO cone and tunnel tests using susceptible *An. gambiae* s.s. were conducted for quality control, while field-collected mosquitoes were used for bio-efficacy monitoring. Molecular species identification was performed using PCR assays, and genomic DNA was extracted from *An. funestus* s.l. and *An. gambiae* s.l. collected from the trials.

All ITNs met WHO bio-efficacy criteria in cone and tunnel tests. *An. gambiae* s.l. predominated in Mwagagala (77%) but was rare in Magu (4%), where *An. funestus* complex was dominant. In Magu, Interceptor® G2 and Veeralin® achieved significantly higher mortality than MAGNet® at 24 h (53% and 45% vs 24%) and 72 h (61% and 53% vs 24%). In contrast, no significant mortality differences were observed among ITNs in Mwagagala. High resistance intensity was detected in both vector groups (mortality <20% in bioassays). In *An. funestus* from Magu, the 4.3 kb-SV resistant genotype was fixed, while G454A-CYP9K1 resistance was highly prevalent (RR: 0.70– 1.00).

Interceptor® G2 and Veeralin® outperformed the pyrethroid-only reference net against *An. funestus* complex but not against predominantly *An. arabiensis* populations.

## Introduction

Malaria continues to present a significant global health challenge, with approximately 282 million cases and 610,000 deaths estimated in 2024, of which 96% occurred in the World Health Organization (WHO) African Region (1). In response, efforts to curb malaria have led to the distribution of 2.3 billion insecticide-treated nets (ITNs) worldwide between 2004 and 2022, with 86% of these delivered to sub-Saharan Africa (2). In 2020 alone, an estimated 229 million ITNs were provided to malaria-endemic regions, including 19.4 million pyrethroid-piperonyl butoxide (PBO) nets (2), and more recently pyrethroid-chlorfenapyr ITNs have begun to be deployed at scale (3). Vector control interventions including ITNs are credited with averting 1.7 billion malaria cases and 10.6 million deaths between 2000 and 2020 (2). However, as ITN distribution has expanded in sub-Saharan Africa, pyrethroid resistance in mosquito vector populations has also surged, undermining ITN effectiveness (4–6).

To combat insecticide resistance, new malaria control tools have been developed. The first of these new ITNs combined a pyrethroid with piperonyl butoxide (PBO), a synergist designed to improve net efficacy against resistant mosquitoes. This class of ITN was first evaluated in cluster randomized control trials (cRCTs) in Tanzania (Olyset^TM^ Plus *vs* Olyset^TM^ net) and Uganda (PermaNet® 3.0, Olyset^TM^ Plus, PermaNet® 2.0 and Olyset ^TM^ net), demonstrating a reduction in malaria prevalence by 44% in the pyrethroid-PBO ITN arm, compared to the standard pyrethroid arm, after one year and by 33% after two years in Tanzania (7), and by 14% after 18 months in Uganda (8). Following this, the WHO recognized the public health value of pyrethroid-PBO ITNs, issuing a conditional recommendation for their use (9). Subsequent commercial investments led to the development of dual-active ingredient (A.I.) ITNs, such as Interceptor® G2, which have undergone WHO Phase I and II trials and shown promising results in areas with pyrethroid resistance. Interceptor® G2 combines alpha-cypermethrin with chlorfenapyr, achieving a 71% mortality rate in resistant mosquitoes in experimental hut trials, compared to 20% for alpha-cypermethrin-only nets (10). A large-scale cRCT in Tanzania found that Interceptor® G2 ITNs reduced malaria infection by 55% and the entomological inoculation rate by 85% after two years of use, compared to pyrethroid-only nets (11). A second cRCT conducted in Benin showed similar results with Interceptor® G2, reducing malaria incidence by 44% compared to pyrethroid-only nets(12). Recently, a new type of bed net coated with chlorfenapyr (PermaNet® Dual) has been developed for malaria control. In Siaya Kenya, an entomological study comparing the efficacy of Interceptor® G2 and PermaNet® Dual reported non-inferiority between these two ITNs (13).

While it is anticipated that these novel dual-A.I. ITNs will provide superior protection from malaria, compared to conventional pyrethroid-only nets; differences in their physical specifications (including location and loading dose of PBO or partner insecticide, the type and content of pyrethroid, wash-fastness and bioavailability of PBO or partner insecticide) mean that their relative performances cannot be assumed to be equivalent in a single study site. Furthermore, differences in vector species composition, behaviour, levels of insecticide resistance intensity and underlying molecular and metabolic resistance mechanisms will also influence LLIN comparative efficacy between malaria endemic areas (14).

The Resilience Against Future Threats through Vector Control (RAFT) programme was a 6-year research consortium, funded by the UK Foreign, Commonwealth and Development Office, which brought together collaborative partners from across sub-Saharan Africa (Institut Pierre Richet, Côte d’Ivoire;; Center for Research in Infectious Diseases, Cameroon;; National Institute for Medical Research, Tanzania), South-East Asia (Mahidol University, Thailand) and the UK (London School of Hygiene and Tropical Medicine, Imperial College London, Malaria Consortium), to generate robust entomological evidence to guide national and global vector-control decision-making.

RAFT consortium partners conducted parallel experimental hut trials in Cameroon, Côte d’Ivoire and Tanzania to assess the impact of different brands of dual-A.I. LLINs on key malaria entomological indicators and their potential for resistance management. Here we report the results from Tanzania.

## Methods

### Study sites

Experimental hut trials were conducted at Welamasonga village in Magu district and at Mwagagala village in Misungwi district; two neighbouring districts along the Lake Zone in Northwestern Tanzania sharing similar environmental conditions and community cultural characteristics. Rainfall ranges from 0.5 to 58.8 mm annually, distributed across two rainy seasons (October–December and March–May), separated by a long dry season (June–August or September) and a shorter dry season (December or January–February). The primary malaria vector species are *An. funestus* sensu stricto (s.s.), *An. arabiensis*, and *An. gambiae* s.s. Species composition varies seasonally, with *An. gambiae* s.l. being more prevalent during the rainy season and *An. funestus* s.l. during the dry season. Most residents are subsistence farmers, relying on rainfall to grow crops like rice, cotton, maize, millet, and legumes, and they raise livestock, including cattle, goats, and sheep. In Misungwi, the primary malaria control measures include universal coverage of ITNs and historical indoor residual spraying (IRS). ITNs were last distributed in 2019, and IRS using Actellic 300CS was last implemented in 2015. Although geographically and culturally alike, the two sites differ in mosquito species composition, with one site having a higher abundance of *Anopheles* (*An*.) *funestus* s.l., providing a useful contrast for evaluating interventions across different vector profiles.

In Magu, the experimental veranda-trap huts are located at 2.5779356° S, 33.1194747° E, in the northern part of Misungwi district Tanzania Initial resistance testing demonstrated low mortality (69%) amongamong mosquitoes exposed to the discriminating concentration (DC; 0.05%) of alpha-cypermethrin in WHO tube assays; mortality increased to 96% after exposure to 10× the DC. By comparison, mortality on permethrin papers was even lower at the DC (0.75%), with 25% mortality for *An. funestus* s.l. and 13% for *An. gambiae* s.l. in Magu (15).

In Mwagagala, the experimental huts are located at 3.0731° S, 32.9888° E, in Mwagagala village, the southern part of Misungwi district. Like Magu, the main vectors in Mwagagala are *An. gambiae* s.s., *An. arabiensis*, and *An. funestus* s.s (11). Previously, high resistance intensity to alpha-cypermethrin and permethrin (<60% mortality) in *An. gambiae* s.l. and *An. funestus* s.l. was reported (16, 17).

### Experimental hut trial design

The study used East African experimental huts with (Mwagagala) and without (Magu) verandas in two sites (six huts in each site). The study was conducted during rain season. Each experimental hut trial was performed according to WHO guidelines, against free flying wild *An. gambiae* s.l. and *An. funestus* s.l.. A total of four PBO/dual-A.I. ITNs (Interceptor® G2, PermaNet® 3.0, Veeralin®, and Olyset^TM^ Plus) were rotated daily to evaluate their efficacy in comparison to a pyrethroid-only net (MaGNet®) (Table 1). An untreated net was included in the rotation to estimate blood feeding inhibition.

**Table 1:** Characteristics of ITNs under evaluation.

| ITN brand | Dose AI/m<sup>2</sup> of netting fabric | Fibre and Denier | Manufacturer |
| --- | --- | --- | --- |
| Olyset™ Plus | 1. Permethrin 800 mg<br>2. PBO 400 mg | 150 polyethylene | Sumitomo chemical Co., Ltd |
| Interceptor® G2 | 1. Alpha-cypermethrin 100 mg<br>2. Chlorfenapyr 200 mg | 100 polyester | BASF Corporation |
| Veeralin® | 1. Alpha-cypermethrin 216mg<br>2. PBO 79 mg | 130 polyethylene | V. K. A. Polymers Pvt. Ltd. |
| PermaNet® 3.0 | 1. Deltamethrin 120 mg<br>2. PBO 800 mg | 75-100 polyester (sides part) and 180 polyethylene (roof part) | Vestergaard Frandsen |
| MaGNet® | 1. Alpha-cypermethrin 216 mg | 150 polyethylene | V. K. A. Polymers Pvt. Ltd. |

For each net brand, we used four new nets, with six 4 x 4 cm holes cut according to WHO guidelines (18). Each experimental hut trial ran for six weeks, utilizing six experimental huts with the treatments specified in Table 1. To account for individual variation in human attractiveness to mosquitoes, sleepers rotated between huts each night, while treatments were rotated weekly in a randomized Latin square design. Over the 36-day trial, each net type was rotated nightly.

Inside each hut, cloth sheets were placed on the floor, and sugar solution was provided in the window traps at night to reduce mosquito mortality. The day after ITN installation, mosquitoes were collected from the hut floor, inside nets, and on walls, ceilings and veranda using standard mouth aspirators. Collected mosquitoes were put in paper cups, labelled with the collection date, hut number, net type, collection area (exit/room/net), and collector initials. All samples were sent to the National Institute for Medical Research (NIMR) insectary for further analysis. Mosquitoes were identified morphologically to the species complex level and classified by physiological status (unfed, freshly fed, semi-gravid, and gravid).

Mosquito mortality was monitored daily for up to 72 hours under standardised insectary conditions of 27 ± 2°C and relative humidity of 70 ± 10%. After the 72-hour holding period, molecular species identification was performed with PCR to distinguish *An. funestus* complex members as either *An. funestus* s.s. or *An. parensis* (19). For *An. gambiae* s.l, PCR was used to identify *An. gambiae* s.s. from *An. arabiensis* (20).

### Chemical analysis of ITNs

New ITNs from each of the five treatment groups, which had not been tested in the huts, were used for chemical analysis. A 30 cm × 30 cm piece of netting was cut from each of five designated locations on one net per brand before trial and stored for chemical analysis as per WHO guidelines (18).

At the conclusion of the experimental hut trials, an equivalent 30 cm × 30 cm sample was taken from same five locations on one of the ITNs tested in the field, making a total of 25 net pieces for chemical analysis post experimental hut trial. All net samples for chemical analysis were wrapped in labelled aluminium foil, sealed in bags, and stored at 4°C before shipment to Centre Wallon de Recherches Agronomiques (CRA-W), Belgium for analysis.

### Bio-efficacy testing and sample handling

In accordance with WHO guidelines (21), bio-efficacy was assessed using WHO cone tests using the susceptible *An. gambiae s.s.* Kisumu strain. From each new, unwashed ITN, three pieces (30 × 30 cm) were cut from each side. One piece per side was designated for chemical analysis, while the remaining two were used for bio-efficacy testing. Each sample was labelled with the net identification number, position, and date, with this information both stapled to the sample and written on its aluminium foil wrapper. Samples were then rolled, wrapped in aluminium foil, and stored at 4–8°C in a refrigerator before and between tests.

### Insecticide Bioassays

Mosquitoes for phenotypic resistance testing were collected using standard mouth aspirators and measured using WHO susceptibility and resistance intensity tests (22). Wild pyrethroid-resistant mosquitoes were collected as adults resting indoor in the houses around the experimental huts in both sites. All blood-fed mosquitoes were kept for three days to allow blood-meal digestion before exposure. Discriminating concentrations of alpha-cypermethrin (0.05%), deltamethrin (0.05%), and permethrin (0.75%) were tested individually, with or without pre-exposure to PBO (4%). Each test consisted of four replicates of 20-25 female mosquitoes (either non-blood fed, or gravid individuals of unknown age), exposed to insecticide-treated papers in WHO tubes for 1 hour, with two replicates of untreated control mosquitoes assayed concurrently. *An. gambiae* s.l. and *An. funestus* s.l. were morphologically identified and tested separately.

If mortality was below 98% at the DC, additional testing was conducted with five and ten times the DCs of alpha-cypermethrin (0.25% and 0.5%), deltamethrin (0.25% and 0.5%), and permethrin (3.75% and 7.5%).

Chlorfenapyr susceptibility of wild *Anopheles* mosquitoes was tested using CDC bottle bioassays (23). For these tests, four replicates of 20-25 female mosquitoes (either non-blood fed or gravid individuals of unknown age) were exposed to 100 μg/ml of chlorfenapyr, coated inside a Wheaton 250 ml bottle, for 1 hour. In all assays, the proportion of knockdown was recorded during exposure, and mortality was recorded at 24, 48, and 72 hours post-exposure. If mortality was below 98% at 100 μg/ml chlorfenapyr, testing was repeated with 200 μg/ml chlorfenapyr.

### Molecular Genotyping of resistance markers in mosquitoes from EHTs

Whole mosquitoes of *A. funestus* s.l. and *A. gambiae* s.l. collected from the experimental hut trials (EHTs) conducted in Magu and Mwagagala, respectively, were used for genomic DNA extraction using the Livak protocol (24). Prior to genotyping, members of the *A. funestus* s.l. group were identified using the cocktail polymerase chain reaction (PCR) assay (25), whereas specimens belonging to the *An. gambiae* complex were identified as *An. gambiae* s.s., *An. coluzzii*, or *An. arabiensis* using the SINE PCR protocol (20, 26).Whole mosquitoes of *A. funestus* s.l. and *A. gambiae* s.l. collected from the experimental hut trials (EHTs) conducted in Magu and Mwagagala, respectively, were used for genomic DNA extraction using the Livak protocol (24). Members of the *A. funestus* s.l. group were identified using the cocktail polymerase chain reaction (PCR) assay (25), whereas specimens belonging to the *An. gambiae* complex was identified as *An. gambiae* s.s., *An. coluzzii*, or *An. arabiensis* using the SINE PCR protocol (20, 26).

Genotyping of 4.3 kb-SV, L119F-GSTe2, CYP6P9a/b resistance marker in *Anopheles funestus* s.ll.

An allele-specific PCR was used to genotype the L119F-GSTe2 mutation in the free flying F0 mosquito population from Magu EHTs, using the protocol previously described (27, 28);. the presence of the CYP6P9a/b_R allele was assessed using PCR-RFLP (29, 30); whereas the 4.3 kb-SV was genotyped using a multiplex PCR Assay recently designed (31). The genotyping of the G454A-CYP9K1 resistance markers was performed using a previously established protocol.

### Genotyping of L995F, L995S, R254K resistance markers in *A. gambiae* s.l

The L995F-*kdr* (formerly *kdr*-west) mutation, the L995S-*kdr* (formerly *kdr*-east) and the R254K-*kdr*, which are involved in the DDT and the pyrethroid resistance, were genotyped in *A. gambiae* s.l. populations collected from the Mwagagala EHTs. All the genotyping was performed using TaqMan or LNA assays (32, 33).

### Outcome measures

The two primary entomological outcomes were:

i. Mortality (proportion of mosquitoes that were dead within 24-h up to 72-h post holding)
ii. Blood-feeding inhibition (reduction in blood-feeding of mosquitoes compared to the control huts).

Secondary endpoints were:

i. Deterrence (reduction in hut entry, relative to the control huts),
ii. Induced exophily (proportion of mosquitoes that exited early and were found in exit traps).

### Data analysis

Proportional outcomes from the bioassays (mortality) and the hut trials (exophily, blood feeding and mortality) were analysed using generalised linear mixed models (GLMMs) with a binomial distribution and a logit link function fitted to the data using the “lme4” package (34) in R (35). For the experimental hut data, net type and hut number were included as fixed effects and the sleepers and day of mosquito collection were added as random effects. Differences in the number of mosquitoes entering the hut were assessed using GLMM with a Poisson distribution. Pairwise comparisons were performed using the “multcomp” package in R (35).

## Results

### Species composition and genotyping of key resistance markers

Two experimental hut trials in Magu and Mwagagala with 72 collection nights were conducted. A total of 3548 mosquitoes were collected across hut locations (veranda-trap, exit-trap, room and inside the net). Among them, 2396 [average 5.5 per night] were malaria vectors (*Anopheles*), and the remaining 1162 [2.7] were *Culex quinquefasciatus*. Of the malaria vectors collected, 374 were collected from Magu site and 2022 were collected from Mwagagala site. For all collection periods, 94.4% [1849/1956] of *An. gambiae* s.l. were collected from Mwagagala, and 5.5% [107/1956] were collected from Magu site; however, fewer *An. funestus* s.l. [39.3%; 173/440] were collected from Mwagagala while Magu collected more [60.7%; 267/440] (Table 2). Of the subsample (n=396) of *An. funestus* complex identified to species-level, 98% were *An. funestus* s.s, the remaining were *An. parensis*.

**Table 2:** *Anopheles* mosquitoes collected in the experimental hut trials per site.

| Outcome: Mosquito density | N | Overall<br>% Mean (95%<br>CI) | Mwagagala site |  | Magu site |  |
| --- | --- | --- | --- | --- | --- | --- |
|  |  |  | N | % Mean (95% CI) | N | % Mean (95% CI) |
| Total night collection per hut | 72 | N/A | 36 | N/A | 36 | N/A |
| Mean mosquito per night | 3548 | 8.3 [7.4 - 9.0] | 2779 | 12.9 [11.6 - 14.2] | 769 | 3.6 [3.2 - 3.9] |
| Mean *Anopheles* vectors | 2396 | 5.5 [4.9 - 6.2] | 2022 | 9.4 [8.3 - 10.4] | 374 | 1.7 [1.5 - 1.9] |
| Total *An. gambiae* s.l | 1956 | 4.5 [3.9 - 5.2] | 1849 | 8.6 [7.6 - 9.6] | 107 | 0.5 [0.4 - 0.6] |
| Total *An. funestus* s.l | 440 | 1.0 [0.9 - 1.1] | 173 | 0.8 [0.7 - 0.9] | 267 | 1.2 [1.0 - 1.4] |
| Total *Culex* species | 1167 | 2.7 [2.4 - 2.9] | 768 | 3.6 [3.1 - 4.0] | 399 | 1.9 [1.6 - 2.2] |

### Genotyping of key resistance markers in *An.funestus* s.s and *An.gambiae* s.s

Among *An. funestus* from Magu, all genotyped specimens were homozygous resistant at the 4.3 kb-SV locus and homozygous susceptible at the L119F-GSTe2 and InDel AA_CYP6P9a loci, irrespective of treatment or survival status. The G454A-CYP9K1 resistant genotype was also common across groups, with RR frequencies ranging from 0.70 to 1.00, although occasional heterozygous and homozygous susceptible individuals were detected in some treatment/survival conditions

For the *An. gambiae* genotyped, 77% were *An. arabiensis* and remaining 23% were *An. gambiae* s.s. All *An. arabiensis* genotyped from Mwgagala, regardless of survival status or treatment condition, were homozygous for the susceptible allele at the R254-*kdr*, L995F-*kdr* and L995S-*kdr* loci (Additional file 1). A strong association (homozygous resistant =100%) was observed between the 4.3 kb structural variant (4.3k-SV) and survival following insecticide exposure in both treatment groups, as evidenced in both live and dead mosquito analysed.

### Mortality rates in experimental huts

Overall, the untreated net hut mortality was less than 5% after the 24 hours holding period. For mortality at 24 hours, no significant difference was observed for PBO/dual-A.I. ITNs (Interceptor® G2 47%, 95% CI [34 - 60], Olyset^TM^ Plus 46%, 95% CI [36 - 56], PermaNet® 3.0 43%, 95% CI [33 - 53] and Veeralin® 51%, 95% CI [41 - 62]), compared to mortality in the pyrethroid-only MaGNet® 36%, 95% CI [26-47] (Table 3). The same trend was observed after the 72 hours holding period (Table 3). When the data were analysed per site of collection, only Interceptor® G2 (53%, p=0.005) and Veeralin® (45%, p=0.048) were superior to the positive control in Magu (Figure 1); while in Mwagagala, none of the dual-A.I. ITNs demonstrated significant superiority to the pyrethroid-only net (Figure 2).

**Figure 1:**
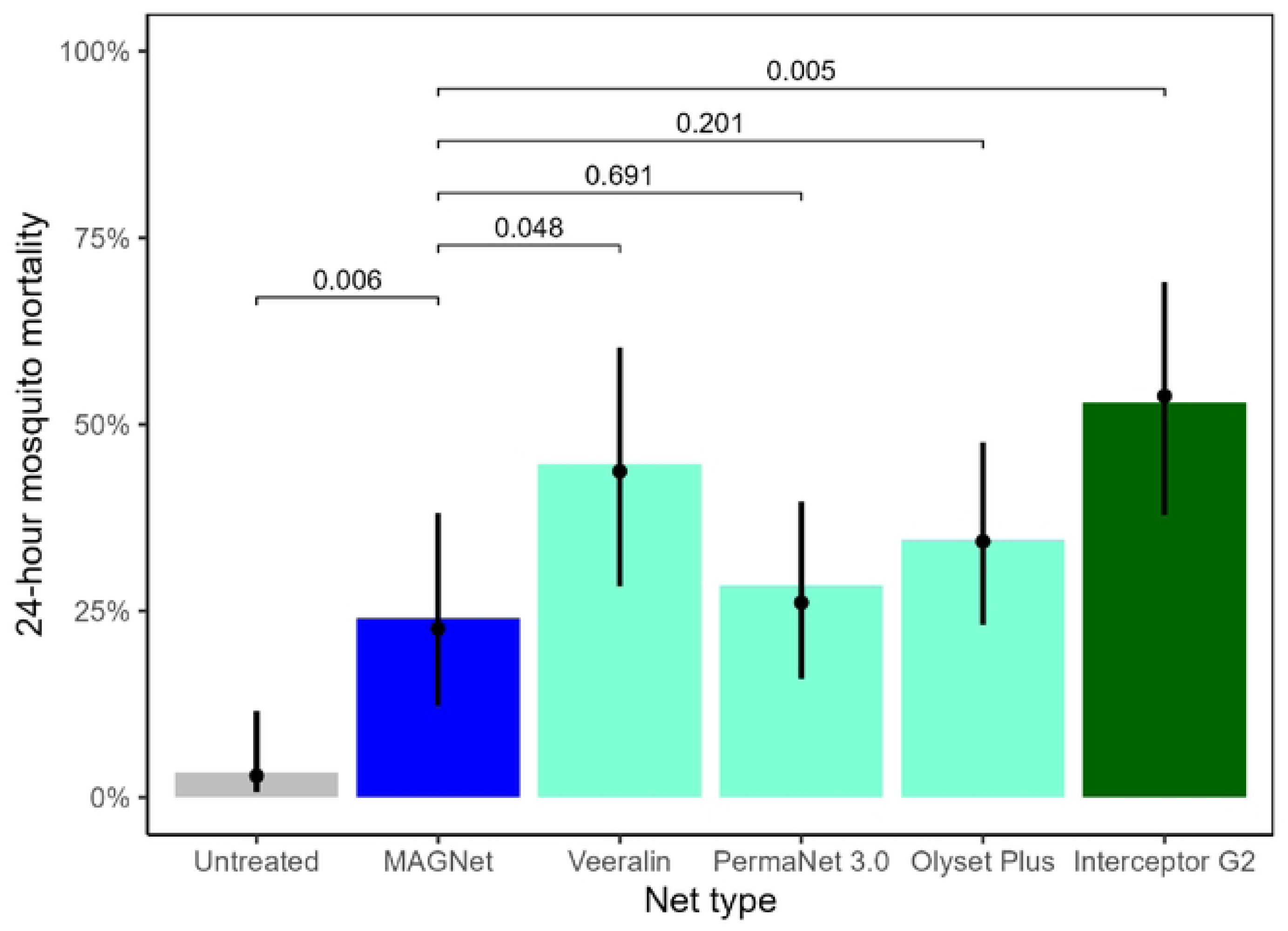
Twenty-four-hour mosquito mortality recorded from Magu experimental hut trial. Bars indicate mean estimates across all huts, with the black points indicating mean estimate generated by the mixed effect logistic regression model, with the vertical lines denoting 95% confidence intervals on this estimate. Bar colours denote the class of net, be it untreated (grey), pyrethroid-only (blue), pyrethroid-PBO (turquoise), or pyrethroid-pyrrole (dark green). Horizontal lines and associated numbers indicate the significance of the difference between each net and the positive control pyrethroid-only net, with values <0.05 assumed to be significantly different.

**Figure 2:**
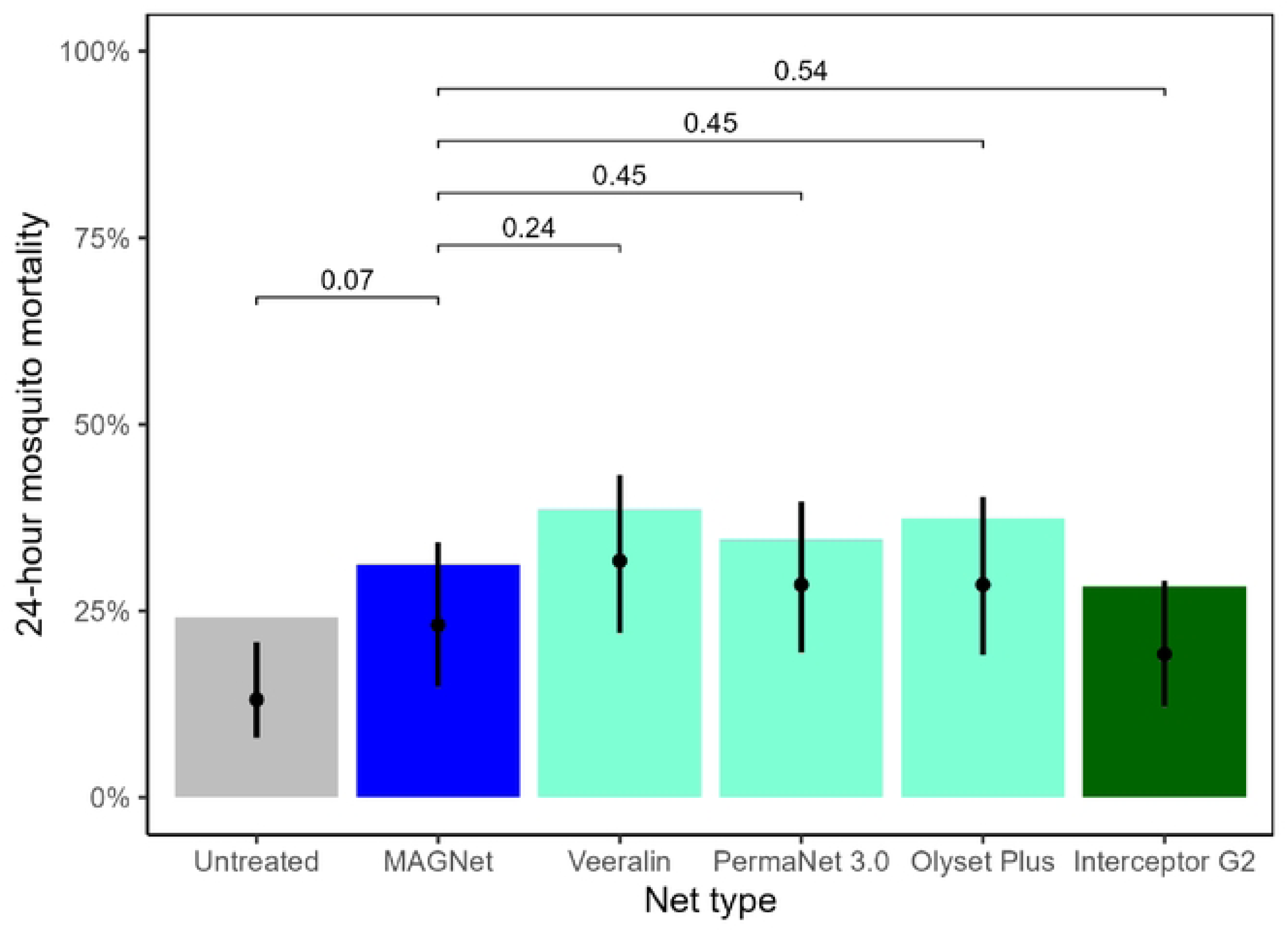
Twenty-four-hour mosquito mortality recorded from Mwagagala experimental hut trial. Bars indicate mean estimates across all huts, with the black points indicating mean estimate generated by the mixed effect logistic regression model, with the vertical lines denoting 95% confidence intervals on this estimate. Bar colours denote the class of net, be it untreated (grey), pyrethroid-only (blue), pyrethroid-PBO (turquoise), or pyrethroid-pyrrole (dark green). Horizontal lines and associated numbers indicate the significance of the difference between each net and the positive control pyrethroid-only net, with values <0.05 assumed to be significantly different.

**Table 3:**
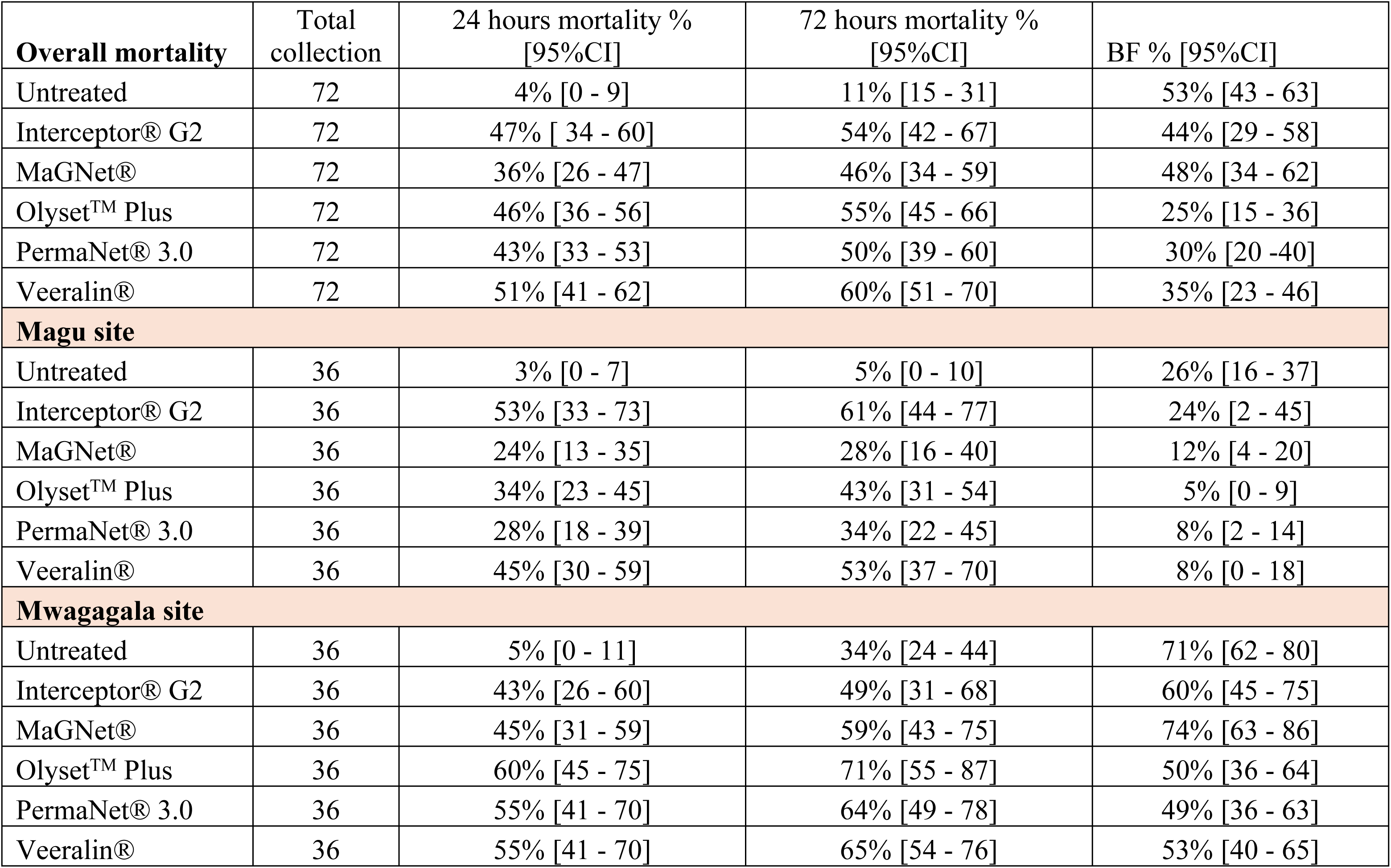
Cumulative mortality for *Anophele*s mosquitoes collected per net per site.

Disaggregating by vector species across both sites, Interceptor® G2 induced 2.24 odds of increased mortality (p=0.057) against *An. funestus* s.l. compared to the reference (MaGNet®). There was no statistically significant difference in the rest of PBO/dual-A.I. ITNs tested compared to the pyrethroid-only net (Table 4).

**Table 4:**
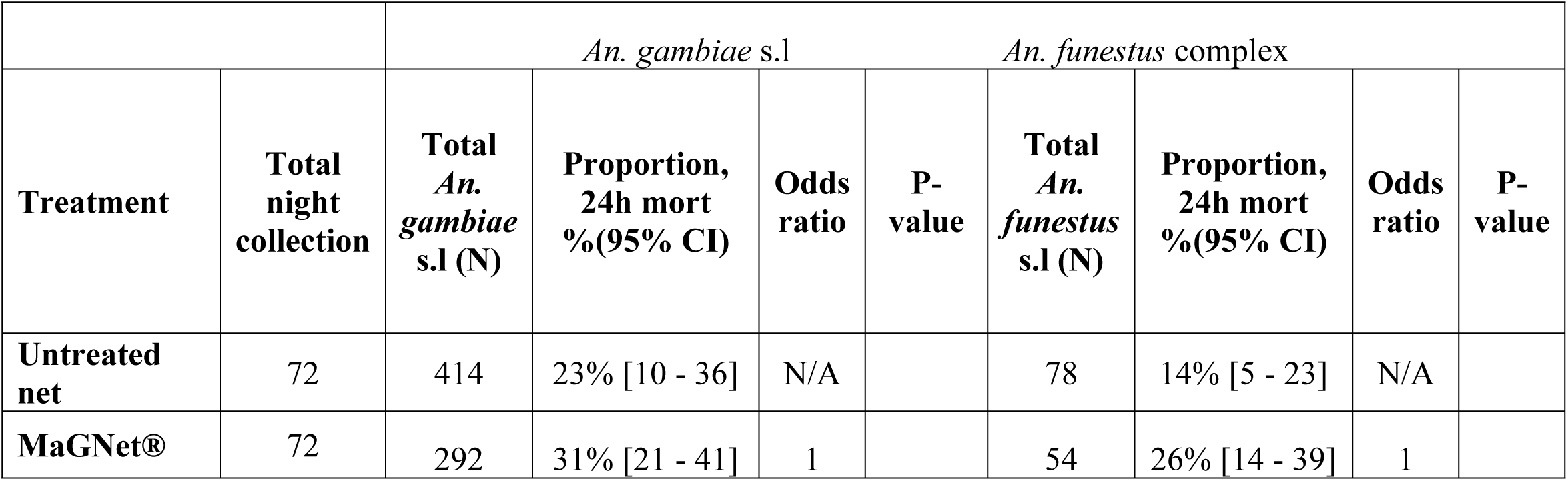
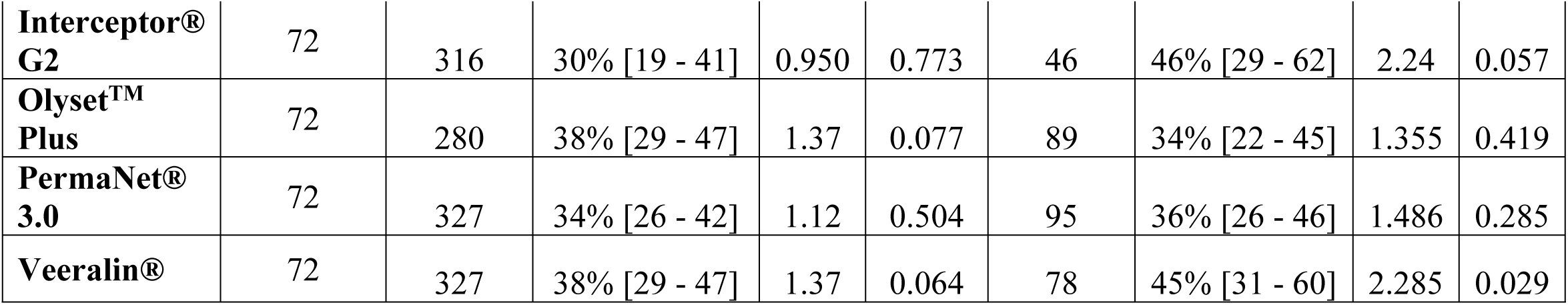
Mortality recorded per species complex per net type.

### Blood feeding

Very low blood feeding was recorded in Magu (Figure 3) compared to Mwagagala (Figure 4). In Mwagagala, only PermaNet® 3.0 significantly reduced the number of mosquitoes that were blood fed (p>0.01) compared to the positive control (MaGNet®), while Interceptor® G2 and Veeralin® were borderline significant (p=0.06) in inhibiting blood feeding in the same site. Nosignificant difference in blood feeding rates was observed in Magu.

**Figure 3:**
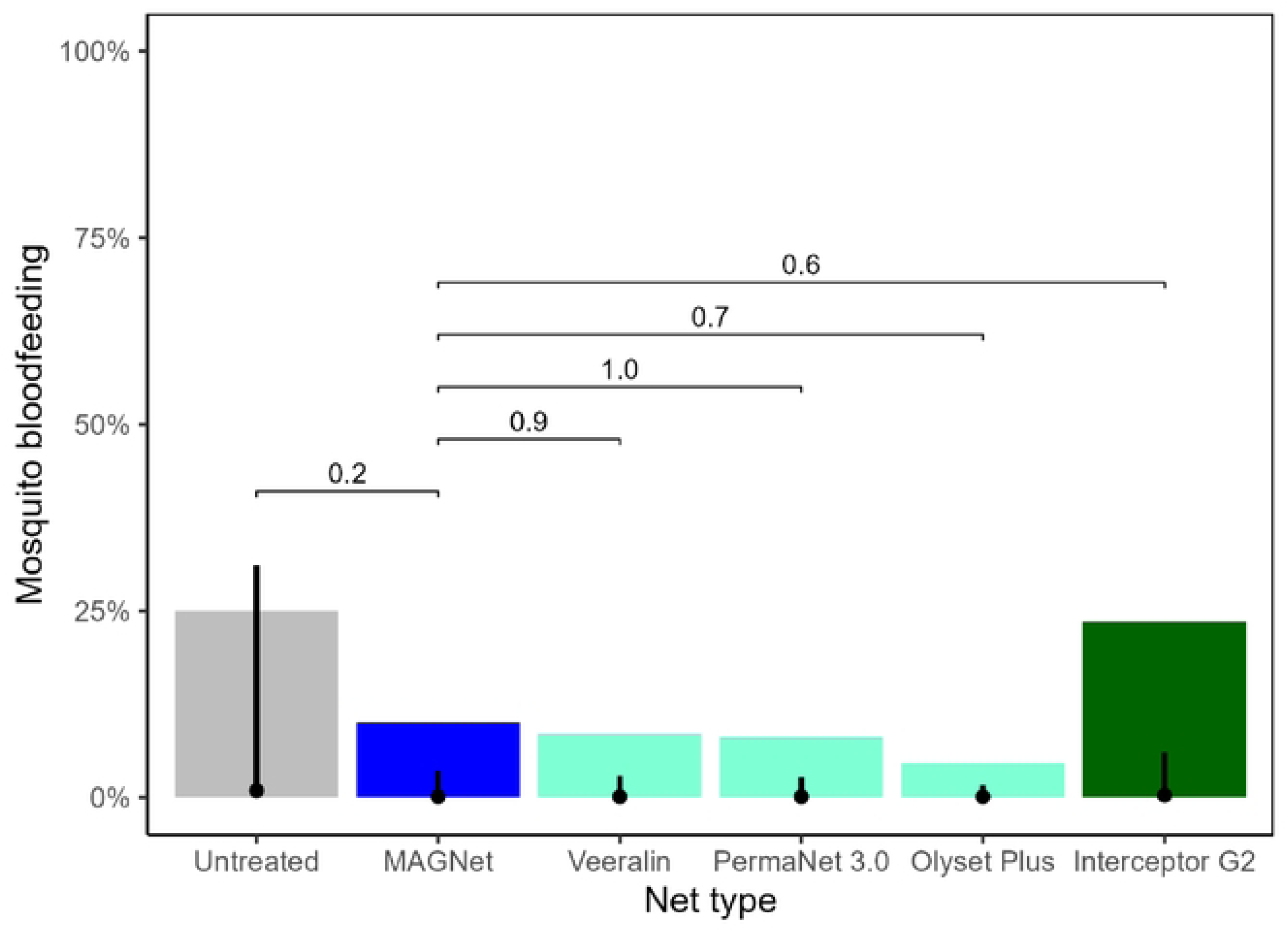
Blood feeding rate observed in Magu.

**Figure 4:**
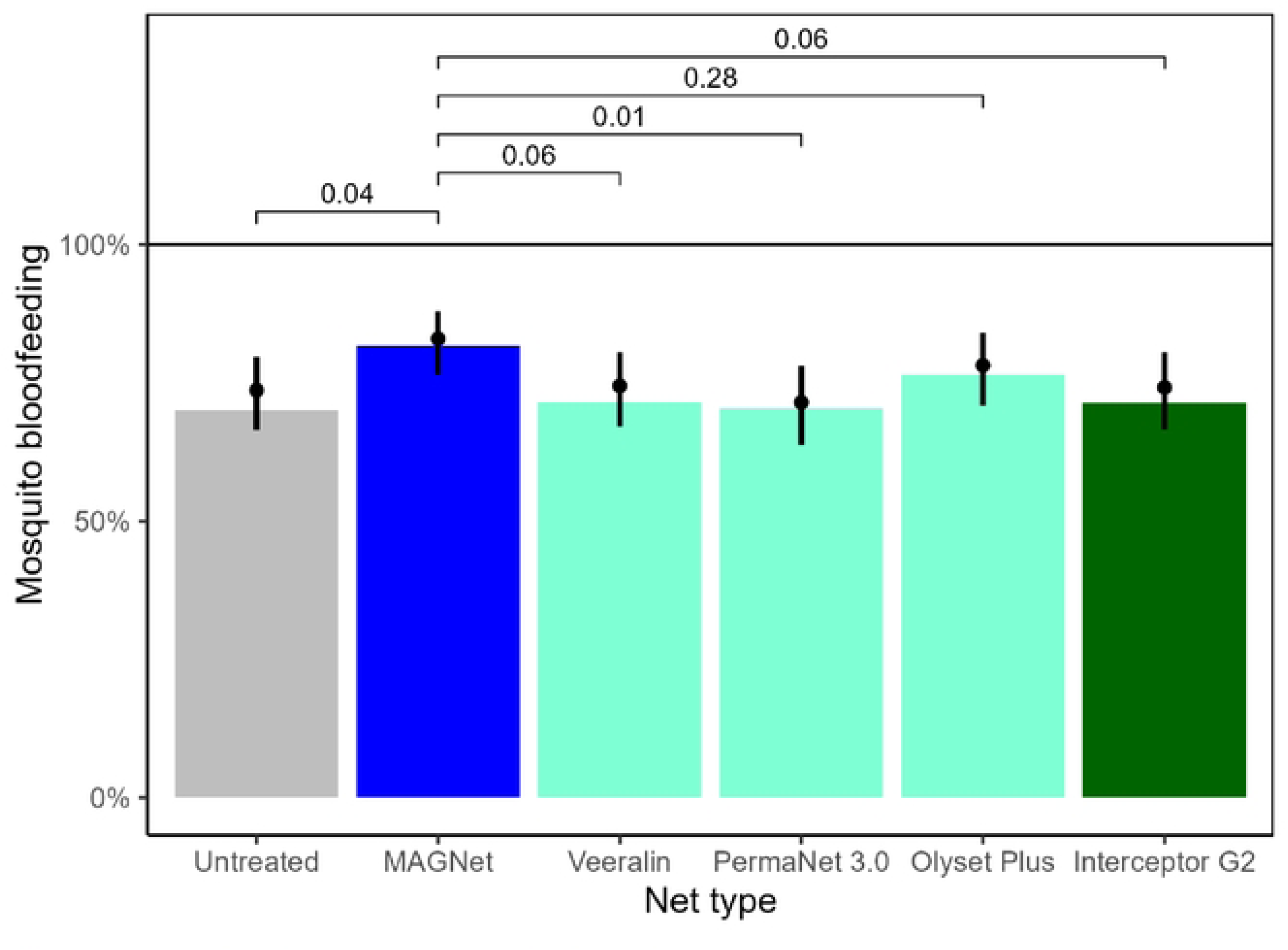
Blood feeding rate observed in Mwagala.

### Exit, deterrence and penetration rate

In all *Anopheles* collected, the exit rate was low for Interceptor® G2 (49%; p=0.002) and Olyset^TM^ Plus (59%; p=0.016) compared to the positive control (MaGNet®; 80%) (Figure 5). No significant deterrence effect was observed in both sites except for Olyset^TM^ Plus, which prevented a significant number of mosquitoes from entering the hut (p=0.02)

**Figure 5:**
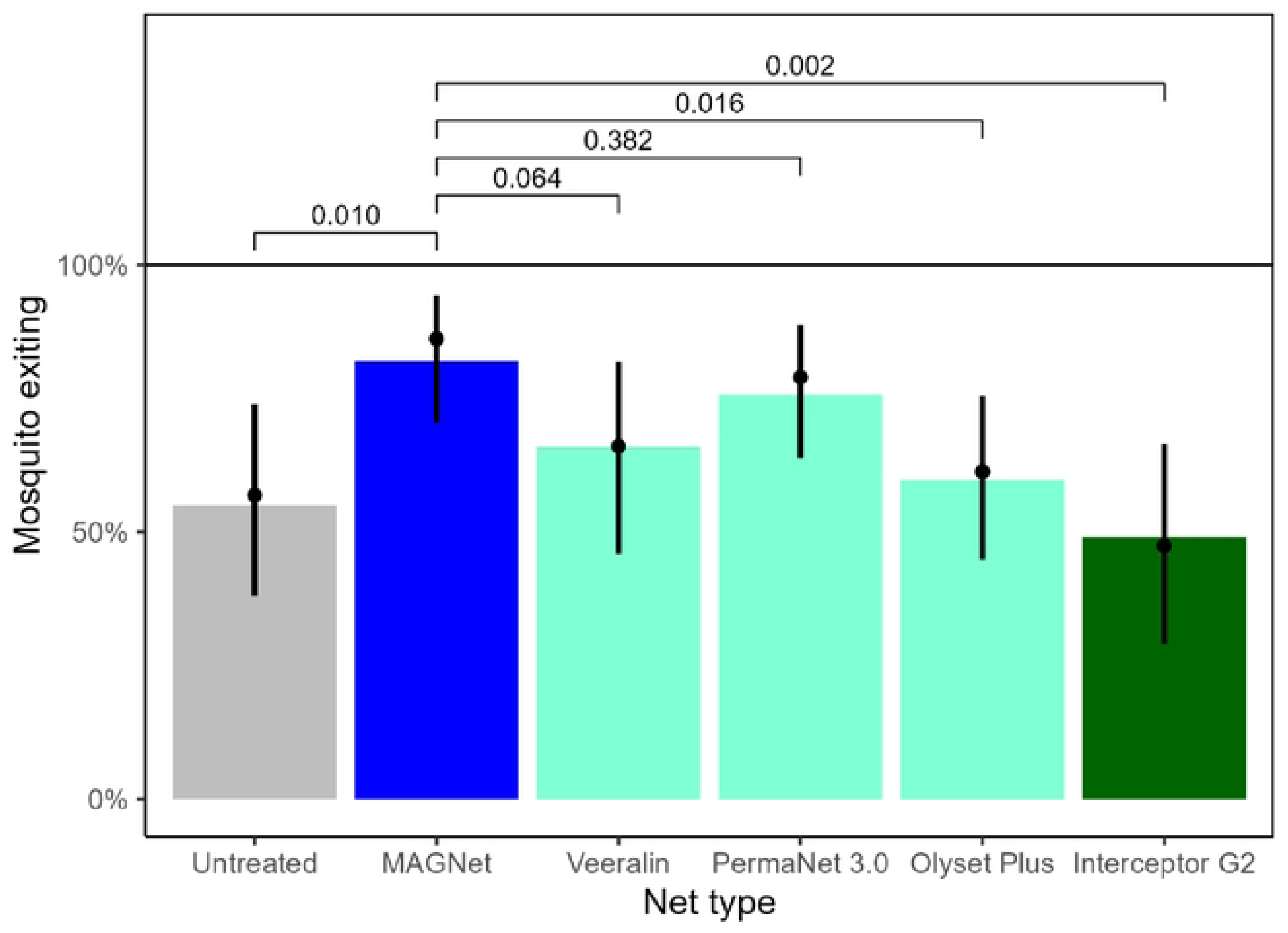
Proportion of mosquitoes exiting the huts. The analysis was performed using mosquitoes collected from Magu site where An. funestus was predominant.

### Mortality in resistance monitoring

High resistance intensity was observed when testing the DC of permethrin (<20% mortality) against *An. gambiae* s.l. and *An. funestus* complex. When the concentration of insecticide was increased by 10 times, mortality remained ≤70% in both species exposed. The same trend was observed when mosquitoes were exposed to either deltamethrin or alpha-cypermethrin. Pre-exposure to PBO did not restore full susceptibility (Additional file 2).

### Chemical content of ITNs and Bio-efficacy results against susceptible strain

Chemical testing of all five samples from each type of net yielded an average target concentration of the insecticide and PBO or partner insecticide for each ITN. All ITNs evaluated met the WHO specifications for insecticide content before and after the trial, with minimal variation of <0.1 in insecticidal content after trial (additional file 5).

All net types evaluated during the experimental huts had mortality >90% against *An. gambiae* s.s Kisumu strain in cone tests and therefore met the WHO bioassay mortality criteria (Additional files 3 & 4).

## Discussion

This study evaluates efficacy of different net brands with different insecticide (Olyset^TM^ Plus, MaGNet®, Interceptor® G2, PermaNet® 3.0, Veeralin®) in area with high pyrethroid resistant malaria vectors. **S**tatistical significance in mortality was recorded against Interceptor® G2 (p=0.005) and Veeralin® (p=0.048) compared to MaGNet® in Magu; however, no statistically significant effect was observed in Mwagagala. This could be explained by the different species collected between both sites. In Magu the predominant species was *An. funestus* s.s. compared to Mwagagala, where most mosquitoes were *An. arabiensis.* From previous studies, Interceptor® G2 has been reported to reduce populations of *An. funestus* s.l. to a greater degree than *An. gambiae* s.l. (14, 15). In Muheza, Tanzania, Veeralin® ITNs were reported to induce higher mortality against *An. funestus* s.l. in experimental hut trials, with low deterrence effect (36). In contrast, in Benin, Interceptor® G2 reduced populations of *An. coluzzii* substantially (6.5-fold higher mortality) compared to untreated nets (37). Similarly, in parallel RAFT experimental hut trials in Cameroon and Côte d’Ivoire against predominantly *An. gambiae* s.s. and *An. coluzzii*, respectively, mortality with Interceptor® G2 was higher than in the present study.

Among the PBO/dual-A.I. ITN assessed, only Olyset^TM^ Plus significantly deterred entry of mosquitoes inside huts. The degree of repellency elicited by PBO/dual-A.I. ITNs will largely reflect the partner pyrethroid, with permethrin inducing excito-repellency to a greater extent than deltamethrin or alpha-cypermethrin. Previous studies from other parts of Tanzania have demonstrated that ITNs had both killing and repellence effect (38). Furthermore, in Kenya, deltamethrin and chlorfenapyr ITNs lacked a deterrence effect on house-entry by malaria vectors (39).

The mean number of *Anopheles* collected was 2.8 times higher than *Culex*. Of the *Anopheles* collected, *An. gambiae* s.l. accounted for 82 % and the remaining 18% were *An. funestus* s.l. More mosquitoes were collected in Mwagagala site than the Magu site; however, there were not statistically significant differences between treatments compared to untreated nets and between species across pooled sites. The species differed slightly in the two sites with *An. gambiae* s.l. being dominant in Mwagagala while *An. funestus* was the major vector species complex in Magu. Of the dead mosquitoes genotyped, the proportion of *An. arabiensis* was higher (77%) than *An. gambiae* s.s. while for *An. funestus* complex collected (N=440), a subsample of 396 mosquitoes was genotyped. Among them, 98% were *An. funestus* s.s and the remaining were *An. parensis*. These results differed from previous reports of higher densities of *An. funestus* s.s. in the southern part of Misungwi district (14). Interestingly, this study found *An. arabiensis* to be more dominant in Mwagagala than in Magu, which may explain differences in vector mortality between sites. Compared to *An. gambiae* s.s.*, An*. *arabiensis* has a greater tendency to feed outdoors (exophagic) and rest outdoors (exophilic), and this may minimize the chances of mosquitoes contacting ITNs. Similar observations have been reported from Tanzania, where a lower effect of dual-A.I. ITNs were evident with *An. arabiensis*, compared to *An. funestus* (15, 40) Overall, a high proportion of female *Anopheles* mosquitoes were collected from the exit trap (64%) in Magu site than in the Mwagagala site (12%). This could be explained by differences in mosquito behaviour between *An. gambiae* s.l. and *An. funestus*.

Resistance intensity was high using the DCs of permethrin, deltamethrin and alpha-cypermethrin against both malaria species (*An. gambiae* s.l. and *An. funestus*) throughout the study period. Pre-exposure to PBO does not restore 100% susceptibility of mosquitoes, except with 10X the DCs, supporting the poorer performances of the pyrethroid-only LLIN and PBO nets during the experimental hut trials against deltamethrin, but full susceptibility was restored after pre-exposure to PBO.

In supplementary assays, after 24 hours of monitoring overall mortality, no statistical significance was observed between PBO/dual-A.I. ITNs and the pyrethroid-only net. Consistent results were observed after 72 hours.

The molecular characterization revealed marked differences in resistance marker profiles between the two study sites. In Mwagagala, the absence of the R254-kdr, L995F-kdr, and L995S-kdr resistance alleles in *An. arabiensis*, irrespective of insecticide exposure or survival status, suggests that target-site *vgsc* mutations are not contributing to the observed phenotypic resistance in this population. This finding indicates that alternative mechanisms, such as metabolic detoxification, are more likely to underline insecticide resistance in this setting. In contrast, *An. funestus* from Magu exhibited a high prevalence of resistance-associated genotypes, particularly the fixation of the 4.3 kb-SV resistant allele and the consistently high frequency of the G454A-*CYP9K1* resistant genotype across treatment and survival groups. The lack of variation in the 4.3 kb-SV, L119F-*GSTe2*, and InDel AA-*CYP6P9a* markers between survivors and non-survivors suggest that these loci alone were not predictive of bioassay outcomes, likely because the resistance alleles are already widespread in the population. Such fixation reduces the ability of genotype–phenotype association analyses to detect significant differences and reflects sustained selection pressure from prolonged insecticide exposure. Continued monitoring of these molecular markers, together with investigations of additional resistance mechanisms, will be important for understanding the evolving resistance landscape and informing vector control strategies.

## Conclusion

Overall, all PBO/dual-A.I. ITNs exhibited higher mosquito mortality compared to the untreated. However, no significant differences in performance were observed for most nets when compared to the reference pyrethroid-only net (MaGNet®, positive control). Notably, Interceptor® G2 and Veeralin® demonstrated superior efficacy against the *anopheles* malaria vectors compared to MaGNet®.

## Data Availability

The data will be available upon request as it is under LSHTM server

## Ethical approval and consent to participate

The study protocol was reviewed and approved by the National Institute for Medical Research (reference No. NIMR/HQ/R.8a/Vol.IX/4166) and the London School of Hygiene and Tropical Medicine (reference No. 28245). Written consent forms were obtained from volunteers (sleepers) who agree to participate in the hut study. Only adults of 18 years or older were recruited, excluding pregnant women. Volunteers were offered daily chemoprophylaxis and the risks of malaria explained (willingness to take part in the study was an inclusion criterion). By sleeping under a mosquito net, they obtain protection not dissimilar to exposure they would normally obtain against mosquitoes in their own home should they use a pyrethroid LLIN. Because an untreated net was used in one arm, all volunteers were monitored each day for signs of fever. Confirmed *falciparum* parasitaemia was treated in nearest health centre.

## Acknowledgements

We extend our sincere gratitude to the local communities for permitting the construction of experimental huts in their areas, and to the project technicians (Patric Hape, Shaban Limbe, and Samora Baltazari Ngero) for their invaluable contributions to mosquito collection, processing, and identification. We also thank the study volunteers for their willingness to participate in the study. We are also grateful to research. Special appreciation goes to the administrative team at the London School of Hygiene & Tropical Medicine—Luke Boddam-Whetham, Maria Bernardez Agrafojo, and Ruth Lorimer at LSHTM whose—for their support enabled the smooth running of activities throughout the project. Our appreciation to manufactures (Sumitomo chemical Co., Ltd, BASF Corporation, V. K. A. Polymers Pvt. Ltd, Vestergaard Frandsen) who willingly provided us with their nets for this study

## Authors contribution

JLM contributed to draft protocol of the study, supervised hut trials and laboratory work, and drafted the manuscript; JL, LAM, MR and SC contributed to draft study protocol, analysis and revised the manuscript; AM was the trial principal investigator, participated in study design, implementation of field activities, data analysis and revised manuscript; JFW and EB contributed to study implementation; TSC, DPD, HCL contribute in the analysis of data and review manuscript; EB contributed to data collection, sorting and storage of mosquitoes; YFT, RW, RN, SA and CW facilitate molecular techniques. All authors read and approved the final manuscript.

